# Nation-wide participation in FIT-based colorectal cancer screening in Denmark during the COVID-19 pandemic: An observational study

**DOI:** 10.1101/2022.08.18.22278786

**Authors:** Tina Bech Olesen, Henry Jensen, Henrik Møller, Jens Winther Jensen, Berit Andersen, Morten Rasmussen

**Author notes:** **Corresponding author** Tina Bech Olesen, MSc, MSc, PhD, Project Manager, Resources & Innovation, The Danish Clinical Quality Program – National Clinical Registries (RKKP), Denmark.

## Abstract

**Background:** Worldwide, most colorectal cancer screening programmes were paused at the start of the COVID-19 pandemic, whilst the Danish faecal immunochemical test (FIT)-based programme continued without pausing. We examined colorectal cancer screening participation and compliance with subsequent colonoscopy in Denmark throughout the pandemic.

**Methods:** We used data from the Danish Colorectal Cancer Screening Database among individuals aged 50-74 years old invited to participate in colorectal cancer screening from 2018-2021 combined with population-wide registries. Using a generalised linear model, we estimated prevalence ratios (PR) and 95% confidence intervals (CI) of colorectal cancer screening participation within 90 days since invitation and compliance with colonoscopy within 60 days since a positive FIT test during the pandemic in comparison with the previous years adjusting for age, month and year of invitation.

**Results:** Altogether, 3,133,947 invitations were sent out to 1,928,725 individuals and there were 94,373 positive FIT tests (in 92,848 individuals) during the study period. Before the pandemic, 60.7% participated in screening within 90 days. A minor reduction in participation was observed at the start of the pandemic (PR=0.95; 95% CI: 0.94-0.96 in pre-lockdown and PR=0.85; 95% CI: 0.85-0.86 in 1^st^ lockdown) corresponding to a participation rate of 54.9% during pre-lockdown and 53.0% during 1st lockdown. This was followed by a 5-10% increased participation in screening corresponding to a participation rate of up to 64.9%. The largest increase in participation was observed among 55-59 year olds, individuals living alone or cohabiting and immigrants. The compliance with colonoscopy within 60 days was 89.9% before the pandemic. A slight reduction was observed during 1^st^ lockdown (PR=0.96; 95% CI: 0.93-0.98), where after it resumed to normal levels.

**Conclusions:** Participation in the Danish FIT-based colorectal cancer screening programme and subsequent compliance to colonoscopy after a positive FIT result was only slightly affected by the COVID-19 pandemic.

**Funding:** The study was funded by the Danish Cancer Society Scientific Committee (grant number R321-A17417) and the Danish regions.

## INTRODUCTION

The COVID-19 pandemic has impacted the society and the healthcare systems worldwide considerably. In efforts to mitigate the impact of the COVID-19 pandemic on the healthcare system and to minimise the spread of the infection, population-wide restrictions (“lockdown”) were imposed worldwide. Large parts of the society were closed down and, within the healthcare system, elective procedures were cancelled or postponed and resources were reallocated to take care of patients in need of hospitalisation because of COVID-19.

As a result of the re-organisations within the healthcare systems, the cancer screening programmes were, in most countries, paused at the start of the pandemic. In Denmark; however, the cancer screening programmes including the faecal immunochemical test (FIT)-based colorectal cancer screening programme using faecal samples obtained at home continued throughout the pandemic. Results from other European countries using FIT-based screening programmes have shown that alterations to the colorectal cancer screening programme at the start of the pandemic led to large reductions in the number of people referred, diagnosed, and treated for colorectal cancer at the start of the pandemic (1, 2) and to reduced participation in screening and screening derived colonoscopy (3) and longer time interval from a positive screening test to colonoscopy (4). A study from Canada also found marked reductions in the colorectal cancer faecal test volumes at the start of the pandemic (5) resulting from a suspension of the FIT-based screening programme. Moreover, it is estimated that the disruptions to the FIT-based colorectal cancer screening programme would result in additional colorectal cancer diagnoses (6). The participation in colorectal cancer screening in Denmark throughout the pandemic has not yet been described; however, one study has shown a 24% reduction in the number of colon cancers diagnosed at the start of the pandemic in Denmark (7) indicating that either the general health-seeking behaviour or the participation in colorectal cancer screening may have changed at the start of the pandemic.

It is well known that social inequities exist across the entire colorectal cancer screening pathway. For example, studies have shown that younger individuals, immigrants, individuals living alone and individuals with a lower income are less likely to participate in colorectal cancer screening (8, 9).Furthermore, the compliance with colonoscopy is lower among older patients and among patients with underlying disease (10), among immigrants and among individuals living alone (9). Moreover, health-seeking behaviour e.g., the number of visits to a general practitioner is associated with participation in colorectal cancer screening (9). A concern is that these social inequities in colorectal cancer screening participation may have been exacerbated during the pandemic.

We examined the colorectal cancer screening participation and compliance with subsequent colonoscopy during the COVID-19 pandemic in Denmark compared with the previous years. Furthermore, we examined whether the participation in colorectal cancer screening and compliance with screen derived colonoscopy during the COVID-19 pandemic differed across population sub-groups.

### Setting

The study was conducted in Denmark, which has a population of approximately 5.8 million inhabitants (11). All residents in Denmark are eligible for tax-supported health care provided by the Danish government. Nationwide population-based registries in Denmark record extensive administrative and medical data of the whole population, which can be linked using the unique personal identifier, that is assigned to all residents at birth or immigration (12, 13).

### The colorectal cancer screening programme

In Denmark, screening for colorectal cancer was implemented in 2014 and is offered free-of-charge every two years to all individuals aged 50-74 years old living in Denmark. The test is a home-based test, which is mailed directly together with an invitation letter to all invitees. The screening procedure is based on a single-sample FIT (OC Sensor (Eiken Chemical Company, Tokyo, Japan)), which can detect invisible amounts of blood in stool samples, which may be associated with bleeding lesions from precancerous adenomas or colorectal cancer at early stages of the disease (14, 15). Non-participants to screening receive a reminder after 6 weeks.

All individuals with a positive FIT test (≥100µg haemoglobin/L faeces) receive an invitation for a colonoscopy with a pre-booked time for appointment within 14 days after the positive screening result. Non-participants to colonoscopy are contacted by the administrative regions.

### The COVID-19 pandemic in Denmark

In Denmark, three main waves of the COVID-19 pandemic have occurred; in the spring of 2020, in the winter of 2020/2021 and again in the winter of 2021/2022 (16).

The pandemic response included population-wide restrictions (lockdowns), COVID-19 testing and COVID-19 vaccination. During the lockdowns large parts of the society were closed down and people were advised to stay at home if possible. Large-scale COVID-19 testing was provided free-of-charge to all inhabitants since May 2020 (17). COVID-19 vaccination began in December 2020 and by March 2022, approximately 81% of the population had received two doses and more than 61% had received three doses of the vaccine (18). The vaccination strategy comprised vaccinating individuals living in nursing homes first, thereafter individuals ≥85 years, then healthcare personnel, thereafter individuals with underlying health conditions and their relatives and finally, individuals were offered the COVID-19 vaccination by decreasing age (75-79 years, 65-74 years, 60-64 years etc.) (19).

### Study population

The study population comprised all invitations in individuals aged 50-74 years old invited to participate in colorectal cancer screening from 1 January 2018 to 30 September 2021, as registered in the Danish Colorectal Cancer Screening Database (20), which contain information on all individuals in Denmark invited to participate in colorectal cancer screening.

To examine participation in colorectal cancer screening, we excluded invitations in individuals who emigrated within 1 year since invitation (N=11,832), invitations in individuals residing in the Faroe Islands or Greenland (N=540), invitations in individuals with an unknown postal address (N=2,621) and registrations of stool samples received before an invitation or reminder was sent out (Supplementary Figure 1A).

To examine compliance with colonoscopy, we included all positive FIT tests from colorectal cancer screening among individuals aged 50-74 years old from 1 January 2018 to 30 September 2021. In all, 146 positive FIT tests were excluded due to low counts across time periods (Supplementary Figure 1B).

### Exposure of interest

The COVID-19 pandemic is the exposure of interest. We defined the different phases of the pandemic in Denmark in accordance with the governmental responses to the COVID-19 pandemic in Denmark, as follows:

- Pre-pandemic period: 1 January 2018 to 31 January 2020
- Pre-lockdown period: 1st February to 10 March 2020
- 1st lockdown: 11 March to 15 April 2020
- 1st re-opening: 16 April to 15 December 2020
- 2nd lockdown: 16 December 2020 to 27 February 2021
- 2nd re-opening: 28 February 2021 to 30 September 2021 (end of inclusion period)

Pre-lockdown and 1^st^ lockdown was termed “the start of the pandemic” in this study.

### Outcome of interests

The two main outcomes of interests were colorectal cancer screening participation within 90 days since invitation and compliance with colonoscopy within 60 days since a positive FIT result.

Further, we evaluated participation within 180 and 365 days since invitation and compliance with colonoscopy within 365 days since a positive FIT result.

### Explanatory variables

The following variables were examined independently: age, sex, ethnicity, cohabitation status, educational level, disposable income and healthcare usage. Age was defined at the date of invitation, as registered in the Danish Colorectal Cancer Screening Database (20). From Statistics Denmark (11), we obtained information on ethnicity, cohabitation status, educational level and level of income. Ethnicity was categorised as Danish descent, Western immigrant, Non-western immigrant and descendants of immigrants. Cohabitation status was categorised as single (i.e. living alone, divorced or not married), co-habiting/co-living, and married (i.e. married or registered partnership) in accordance with Statistics Denmark (11). Education level was defined in accordance with the International Standard Classification of Education (ISCED) of the United Nations Education, Scientific and Cultural Organization (UNESCO) into short (ISCED level 1-1), medium (ISCED level 3-4) and long (ISCED level 5-8) (11). Income was defined as official disposable income depreciated to 2015 level and categorised into five quintiles. To indicate the level of healthcare use by each patient, we counted the total number of contacts to general practitioners, private practising medical specialists, physiotherapists, and chiropractors in the year for invitation as registered in the Danish National Health Service Register (21), which contain information on visits to primary healthcare (e.g., general practitioners and medical specialists) in Denmark since 1990. We categorised healthcare usage as rare (0-3 visits per year), low (4-6 visits per year), average (7-11 visits per year), high (12-18 visits per year) and frequent (≥19 visits per years).

### Statistical analyses

We examined characteristics of persons invited to participate in colorectal cancer screening and characteristics of persons with a positive FIT test during the study period. Thereafter, we examined the participation in colorectal cancer screening within 90, 180 and 365 days since invitation overall and stratifying by the explanatory variables per month and during the pandemic phases. Similarly, we examined compliance with colonoscopy within 60 and 365 days since a positive FIT test overall and stratifying by the explanatory variables per month and during the pandemic phases. We also examined the median number of days and interdecentile interval (IDI) from invitation to participation overall and during the different phases of the pandemic.

Using a generalised linear model (GLM) with log link for the Poisson family with robust standard errors (SE), we estimated prevalence ratios (PR) and 95% confidence intervals (CI) of participation in colorectal cancer screening within 90, 180 and 365 days since invitation among persons invited to participate in colorectal cancer screening and compliance with colonoscopy within 60 and 365 days since a positive FIT test during the different phases of the pandemic overall and stratifying by the explanatory variables. Firstly, we calculated unadjusted analyses. Thereafter, the analyses were adjusted for month of invitation to allow for seasonality and year of invitation to take into account the annual change in colorectal cancer screening participation. Finally, the analyses were adjusted for age to take into account the effect of age on the other explanatory variables.

We performed a sensitivity analyses to take into account an IT-error that occurred in the spring of 2020 resulting in a reduction in the number of invitations sent out in week 11-14 2020 (Central Denmark Region in week 11-14 2020, Northern Denmark Region in week 12-14 2020 and the rest of Denmark in week 13-14). The error meant, that only individuals entering or leaving the screening programme were invited. Thus, during the period with the IT-error only 50 years olds entering the programme, individuals entering the country from abroad, and 73 to 74 year olds leaving the programme were invited. We re-ran the analyses for this by introducing a dummy variable expressing the IT-error in the GLM model.

All analyses were conducted using STATA version 17.0.

### Ethical considerations

The study is registered at the Central Denmark Region’s register of research projects (journal number 1-16-02-381-20). Patient consent is not required by Danish law for register-based studies.

## RESULTS

Altogether, 3,133,947 invitations were sent out to 1,928,725 individuals during the study period. Among those 50.5% were women and the median age was 60 years (IQI=54-67), the majority were of Danish descent (91.4%), most were married (59.4%) and 56% had a short educational level. The distribution of the descriptive characteristics was similar across the pandemic phases (Table 1).

**Table 1.** Baseline characteristics of people invited to participate in colorectal cancer screening in Denmark from 2018 to 2021.

|  | Pre-pandemic<br>(01Jan2018-31Jan2020) | Pre-lockdown<br>(01Feb2020-10Mar2020) | 1st lockdown<br>(11Mar2020-15Apr2020) | 1st re-opening<br>(16Apr2020-15Dec2020) | 2nd lockdown<br>(16Dec2020-27Feb2021) | 2nd re-opening<br>(28Feb2021-30Sep2021) | Total |
| --- | --- | --- | --- | --- | --- | --- | --- |
|  | N (%) | N (%) | N (%) | N (%) | N (%) | N (%) | N (%) |
| **Total** | 1804770 (57.6) | 104892 (3.3) | 40146 (1.3) | 562124 (17.9) | 176469 (5.6) | 445546 (14.2) | 3133947 (100.0) |
| **Sex** |  |  |  |  |  |  |  |
| Men | 893009 (49.5) | 52523 (50.1) | 19285 (48.0) | 276731 (49.2) | 87667 (49.7) | 218713 (49.1) | 1547928 (49.4) |
| Women | 911761 (50.5) | 52369 (49.9) | 20861 (52.0) | 285393 (50.8) | 88802 (50.3) | 226833 (50.9) | 1586019 (50.6) |
| **Age at invitation** |  |  |  |  |  |  |  |
| 50-54 years | 478804 (26.5) | 14982 (14.3) | 7322 (18.2) | 127306 (22.6) | 49217 (27.9) | 138988 (31.2) | 816619 (26.1) |
| 55-59 years | 375309 (20.8) | 38988 (37.2) | 5739 (14.3) | 150615 (26.8) | 32217 (18.3) | 61318 (13.8) | 664186 (21.2) |
| 60-64 years | 332766 (18.4) | 18089 (17.2) | 2929 (7.3) | 101925 (18.1) | 33503 (19.0) | 86806 (19.5) | 576018 (18.4) |
| 65-69 years | 308725 (17.1) | 16334 (15.6) | 2859 (7.1) | 92026 (16.4) | 30398 (17.2) | 79821 (17.9) | 530163 (16.9) |
| 70-74 years | 309166 (17.1) | 16499 (15.7) | 21297 (53.0) | 90252 (16.1) | 31134 (17.6) | 78613 (17.6) | 546961 (17.5) |
| Median (IQR) | 60 (54; 67) | 59 (55; 66) | 72 (56; 75) | 60 (55; 67) | 60 (54; 67) | 61 (54; 67) | 60 (54; 67) |
| **Ethnicity** |  |  |  |  |  |  |  |
| Danish descent | 1651658 (91.6) | 95490 (91.1) | 36665 (91.4) | 512355 (91.2) | 150194 (91.0) | 366141 (91.2) | 2812503 (91.4) |
| Descendant of immigrant | 2932 (0.2) | 169 (0.2) | 65 (0.2) | 987 (0.2) | 284 (0.2) | 617 (0.2) | 5054 (0.2) |
| Western Immigrant | 48061 (2.7) | 2871 (2.7) | 1164 (2.9) | 15877 (2.8) | 4642 (2.8) | 10902 (2.7) | 83517 (2.7) |
| Non-western immigrant | 99730 (5.5) | 6334 (6.0) | 2238 (5.6) | 32755 (5.8) | 9939 (6.0) | 23620 (5.9) | 174616 (5.7) |
| **Cohabitation status** |  |  |  |  |  |  |  |
| Single | 567436 (31.5) | 34149 (32.6) | 14487 (36.1) | 177512 (31.6) | 51320 (31.1) | 120737 (30.1) | 965641 (31.4) |
| Co-habiting/co-living | 163186 (9.1) | 9679 (9.2) | 2758 (6.9) | 53372 (9.5) | 15775 (9.6) | 37404 (9.3) | 282174 (9.2) |
| Married/registered partner | 1071632 (59.5) | 61036 (58.2) | 22887 (57.0) | 331090 (58.9) | 97964 (59.4) | 243139 (60.6) | 1827748 (59.4) |
| **Educational level (ISCED)** |  |  |  |  |  |  |  |
| ISCED15 level 1-2 | 976876 (55.1) | 58698 (57.0) | 18575 (47.2) | 316639 (57.3) | 98365 (56.8) | 250198 (57.2) | 1719351 (55.9) |
| ISCED15 level 3-4 | 597535 (33.7) | 33332 (32.3) | 15297 (38.8) | 177249 (32.1) | 55821 (32.2) | 139785 (32.0) | 1019019 (33.1) |
| ISCED15 level 5-8 | 198531 (11.2) | 11011 (10.7) | 5513 (14.0) | 58592 (10.6) | 18970 (11.0) | 47211 (10.8) | 339828 (11.0) |
| **Disposable income** |  |  |  |  |  |  |  |
| Lowest quintile | 353612 (19.6) | 18722 (17.9) | 9939 (24.8) | 97358 (17.4) | 29966 (17.0) | 74581 (16.8) | 584178 (18.7) |
| Second quintile | 358955 (19.9) | 20579 (19.7) | 9583 (23.9) | 105809 (18.9) | 33736 (19.2) | 83982 (18.9) | 612644 (19.6) |
| Third quintile | 366888 (20.3) | 21023 (20.1) | 7332 (18.3) | 114154 (20.4) | 33347 (18.9) | 82529 (18.6) | 625273 (20.0) |
| Fourth quintile | 366219 (20.3) | 21767 (20.8) | 6614 (16.5) | 119544 (21.3) | 36705 (20.8) | 94378 (21.2) | 645227 (20.6) |
| Highest quintile | 358608 (19.9) | 22588 (21.6) | 6644 (16.6) | 123676 (22.1) | 42412 (24.1) | 108662 (24.5) | 662590 (21.2) |
| **Healthcare usage** |  |  |  |  |  |  |  |
| Rare | 386510 (21.4) | 23944 (22.8) | 7293 (18.2) | 125799 (22.4) | 37849 (21.4) | 95024 (21.3) | 676419 (21.6) |
| Low | 338179 (18.7) | 18944 (18.1) | 6054 (15.1) | 103813 (18.5) | 32147 (18.2) | 80134 (18.0) | 579271 (18.5) |
| Average | 401358 (22.2) | 23271 (22.2) | 8569 (21.3) | 125600 (22.3) | 39176 (22.2) | 98833 (22.2) | 696807 (22.2) |
| High | 333529 (18.5) | 19230 (18.3) | 8343 (20.8) | 104646 (18.6) | 33618 (19.1) | 85706 (19.2) | 585072 (18.7) |
| Frequent | 345194 (19.1) | 19503 (18.6) | 9887 (24.6) | 102266 (18.2) | 33679 (19.1) | 85849 (19.3) | 596378 (19.0) |
| **Time from invitation to participation, median (IDI)** | 28 (16; 72) | 25 (15; 78) | 32 (16; 64) | 28 (16; 66) | 24 (16; 63) | 25 (16; 59) | 28 (16; 70) |

IQI = interquartile interval; IDI = interdecile interval; ISCED=International Standard Classification of Education

### Participation during the COVID-19 pandemic

Before the pandemic, 60.7% participated in colorectal cancer screening within 90 days since invitation (Figure 1 and Supplementary Table 1). The results were similar extending the length of follow-up time to 180 and 365 days (data not shown).

**Figure 1.**
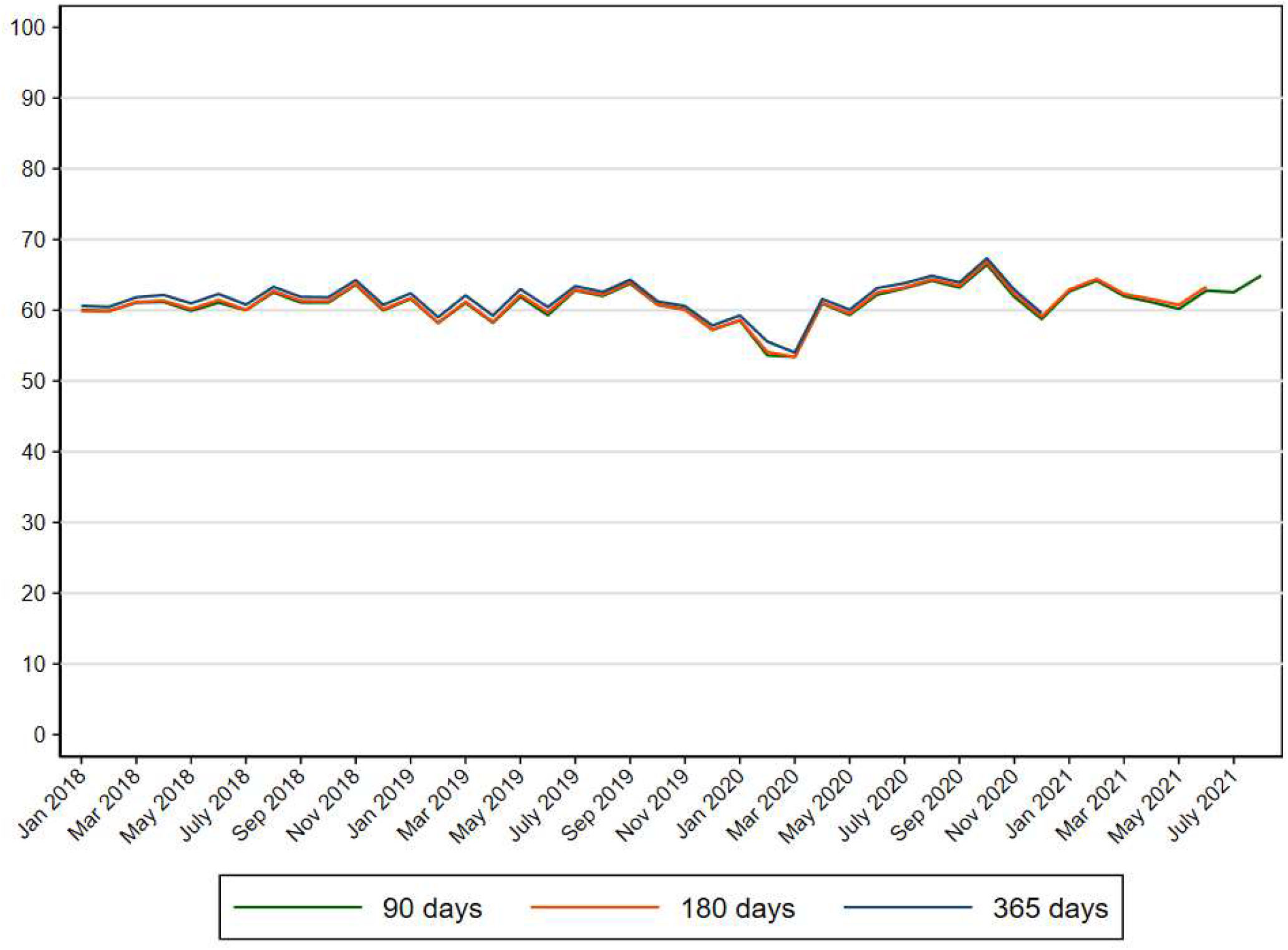
Participation in colorectal cancer screening (%) in Denmark within 90, 180 and 365 days since invitation from 2018 to 2021.

A reduction in screening participation within 90 days occurred during February and March 2020 (Figure 1) reflected in a prevalence ratio (PR) of 0.95 (95% CI: 0.94-0.96) during pre-lockdown and a PR of 0.85 (95% CI: 0.85-0.86) during 1^st^ lockdown (Table 2). This reduction corresponded to an overall participation rate of 54.9% during pre-lockdown and 53.3% during 1^st^ lockdown (Supplementary Table 1). Subsequently, an increase in screening participation was observed (Figure 1) reflected in overall PRs of 1.04 (95% CI: 1.04-1.05), 1.09 (95% CI: 1.09-1.10) and 1.11 (95% CI: 1.10-1.12) during 1^st^ re-opening, 2^nd^ lockdown and 2^nd^ re-opening, respectively (Table 2). These increases corresponded to participation rates of 62.4% during 1^st^ re-opening, 63.0% during 2^nd^ lockdown and 64.9% during 2^nd^ re-opening (Supplementary Table 1). These estimates were similar when extending the length of follow-up time to 180 and 365 days (data not shown).

**Table 2.** Prevalence ratios (PR) and 95% confidence intervals (CI) of participation in colorectal cancer screening within 90 days since invitation in Denmark 2018-2021.

|  |  | Pre-pandemic<br>(01Jan2018-31Jan2020) |  | Pre-lockdown<br>(01Feb2020-10Mar2020) |  | 1st lockdown<br>(11Mar2020-15Apr2020) |  | 1st re-opening<br>(16Apr2020-15Dec2020) |  | 2nd lockdown<br>(16Dec2020-27Feb2021) |  | 2nd re-opening<br>(28Feb2021-30Sep2021) |  |
| --- | --- | --- | --- | --- | --- | --- | --- | --- | --- | --- | --- | --- | --- |
|  | N | N=180477 |  | N=104892 |  | N=40146 |  | N=562124 |  | N=176469 |  | N=445546 |  |
|  |  | PR | [95%CI] | PR | [95%CI] | PR | [95%CI] | PR | [95%CI] | PR | [95%CI] | PR | [95%CI] |
| **Overall** | 3133947 | 1.00 | - | 0.95 | [0.94; 0.96] | 0.85 | [0.85; 0.86] | 1.04 | [1.04; 1.05] | 1.09 | [1.09; 1.10] | 1.11 | [1.10; 1.12] |
| **Sex** |  |  |  |  |  |  |  |  |  |  |  |  |  |
| Men | 1547928 | 1.00 | - | 0.88 | [0.88; 0.89] | 0.91 | [0.89; 0.92] | 1.03 | [1.02; 1.03] | 1.04 | [1.04; 1.05] | 1.07 | [1.07; 1.08] |
| Women | 1586019 | 1.00 | - | 0.92 | [0.92; 0.93] | 0.84 | [0.83; 0.85] | 1.03 | [1.02; 1.03] | 1.03 | [1.03; 1.04] | 1.06 | [1.06; 1.07] |
| **Age at invitation** |  |  |  |  |  |  |  |  |  |  |  |  |  |
| 50-54 years | 816619 | 1.00 | - | 0.89 | [0.87; 0.91] | 1.14 | [1.12; 1.17] | 1.00 | [0.99; 1.01] | 1.04 | [1.03; 1.06] | 1.16 | [1.14; 1.18] |
| 55-59 years | 664186 | 1.00 | - | 0.93 | [0.91; 0.94] | 1.09 | [1.06; 1.11] | 1.11 | [1.10; 1.12] | 1.24 | [1.22; 1.25] | 1.18 | [1.16; 1.19] |
| 60-64 years | 576018 | 1.00 | - | 0.94 | [0.93; 0.96] | 1.03 | [1.00; 1.06] | 1.03 | [1.02; 1.04] | 1.08 | [1.07; 1.10] | 1.09 | [1.07; 1.10] |
| 65-69 years | 530163 | 1.00 | - | 0.96 | [0.95; 0.97] | 0.99 | [0.96; 1.02] | 1.02 | [1.01; 1.03] | 1.07 | [1.06; 1.09] | 1.07 | [1.06; 1.08] |
| 70-74 years | 546961 | 1.00 | - | 1.01 | [1.00; 1.03] | 0.77 | [0.76; 0.79] | 1.04 | [1.03; 1.05] | 1.09 | [1.07; 1.10] | 1.10 | [1.08; 1.11] |
| **Ethnicity** |  |  |  |  |  |  |  |  |  |  |  |  |  |
| Danish descent | 2812503 | 1.00 | - | 0.95 | [0.94; 0.95] | 0.85 | [0.84; 0.86] | 1.04 | [1.04; 1.05] | 1.09 | [1.08; 1.10] | 1.11 | [1.10; 1.12] |
| Descendant of immigrant | 5054 | 1.00 | - | 0.95 | [0.79; 1.14] | 0.83 | [0.62; 1.11] | 1.04 | [0.93; 1.17] | 1.08 | [0.91; 1.28] | 1.12 | [0.94; 1.32] |
| Western Immigrant | 83517 | 1.00 | - | 0.95 | [0.91; 1.00] | 0.88 | [0.82; 0.94] | 1.08 | [1.05; 1.11] | 1.16 | [1.11; 1.21] | 1.20 | [1.15; 1.25] |
| Non-western immigrant | 174616 | 1.00 | - | 0.97 | [0.94; 1.00] | 1.00 | [0.95; 1.05] | 1.11 | [1.09; 1.13] | 1.18 | [1.15; 1.22] | 1.24 | [1.20; 1.28] |
| **Cohabitation status** |  |  |  |  |  |  |  |  |  |  |  |  |  |
| Single | 965641 | 1.00 | - | 0.94 | [0.93; 0.95] | 0.85 | [0.83; 0.87] | 1.06 | [1.05; 1.07] | 1.12 | [1.11; 1.14] | 1.16 | [1.14; 1.17] |
| Cohabiting | 282174 | 1.00 | - | 0.95 | [0.92; 0.97] | 0.97 | [0.93; 1.00] | 1.05 | [1.04; 1.07] | 1.12 | [1.09; 1.14] | 1.14 | [1.12; 1.17] |
| Married | 1827748 | 1.00 | - | 0.95 | [0.95; 0.96] | 0.86 | [0.85; 0.87] | 1.04 | [1.03; 1.04] | 1.08 | [1.07; 1.09] | 1.09 | [1.09; 1.10] |
| **Educational level (ISCED)** |  |  |  |  |  |  |  |  |  |  |  |  |  |
| ISCED15 level 1-2 | 1719351 | 1.00 | - | 0.95 | [0.94; 0.95] | 0.91 | [0.90; 0.92] | 1.05 | [1.04; 1.05] | 1.09 | [1.08; 1.10] | 1.12 | [1.11; 1.13] |
| ISCED15 level 3-4 | 1019019 | 1.00 | - | 0.96 | [0.94; 0.97] | 0.80 | [0.79; 0.82] | 1.04 | [1.03; 1.04] | 1.09 | [1.08; 1.10] | 1.10 | [1.09; 1.11] |
| ISCED15 level 5-8 | 339828 | 1.00 | - | 0.95 | [0.94; 0.97] | 0.83 | [0.81; 0.85] | 1.05 | [1.04; 1.06] | 1.11 | [1.09; 1.13] | 1.12 | [1.10; 1.13] |
| **Disposable income** |  |  |  |  |  |  |  |  |  |  |  |  |  |
| Lowest quintile | 584178 | 1.00 | - | 0.95 | [0.94; 0.97] | 0.73 | [0.71; 0.75] | 1.04 | [1.03; 1.05] | 1.08 | [1.06; 1.10] | 1.11 | [1.09; 1.12] |
| Second quintile | 612644 | 1.00 | - | 0.96 | [0.95; 0.98] | 0.78 | [0.76; 0.79] | 1.05 | [1.04; 1.06] | 1.10 | [1.08; 1.11] | 1.13 | [1.11; 1.14] |
| Third quintile | 625273 | 1.00 | - | 0.95 | [0.94; 0.97] | 0.86 | [0.84; 0.88] | 1.05 | [1.04; 1.06] | 1.08 | [1.07; 1.10] | 1.11 | [1.10; 1.12] |
| Fourth quintile | 645227 | 1.00 | - | 0.95 | [0.94; 0.96] | 0.94 | [0.92; 0.96] | 1.04 | [1.04; 1.05] | 1.08 | [1.07; 1.10] | 1.10 | [1.09; 1.11] |
| Highest quintile | 662590 | 1.00 | - | 0.93 | [0.92; 0.94] | 0.98 | [0.96; 1.00] | 1.04 | [1.03; 1.05] | 1.10 | [1.09; 1.11] | 1.10 | [1.09; 1.11] |
| **Healthcare usage** |  |  |  |  |  |  |  |  |  |  |  |  |  |
| Rare | 676419 | 1.00 | - | 0.92 | [0.90; 0.93] | 0.93 | [0.90; 0.95] | 1.04 | [1.03; 1.05] | 1.10 | [1.08; 1.12] | 1.11 | [1.09; 1.12] |
| Low | 579271 | 1.00 | - | 0.95 | [0.94; 0.97] | 0.90 | [0.88; 0.92] | 1.05 | [1.04; 1.06] | 1.10 | [1.08; 1.11] | 1.11 | [1.10; 1.13] |
| Average | 696807 | 1.00 | - | 0.95 | [0.94; 0.97] | 0.86 | [0.85; 0.88] | 1.05 | [1.04; 1.06] | 1.09 | [1.08; 1.10] | 1.11 | [1.10; 1.12] |
| High | 585072 | 1.00 | - | 0.97 | [0.95; 0.98] | 0.84 | [0.82; 0.86] | 1.05 | [1.04; 1.05] | 1.09 | [1.08; 1.11] | 1.12 | [1.10; 1.13] |
| Frequent | 596378 | 1.00 | - | 0.97 | [0.95; 0.98] | 0.81 | [0.79; 0.83] | 1.04 | [1.03; 1.05] | 1.09 | [1.08; 1.10] | 1.11 | [1.10; 1.12] |

\* Adjusted for year, month and age at invitation; PR=prevalence ratio; CI=confidence interval; ISCED=International Standard Classification of Education

### Participation during the COVID-19 pandemic according to socio-economic variables

Throughout the study period, the participation in colorectal cancer screening was lowest among the youngest age group, among men, among immigrants, among individuals living alone or cohabiting, among individuals with a low educational level, a low income and among individuals who rarely use the healthcare system (Supplementary Figure 2). During 1^st^ lockdown, women, 70-74 year olds and individuals with a low income had the lowest participation in screening (Table 2). From 1^st^ reopening and onwards, the largest relative increases in participation was observed among 55-59 year olds, among immigrants and among individuals living alone or co-habiting (Table 2).

### Participation among first-time invitees

Altogether, 8.8% (N=276,495) of the study population were first-time invitees. The median age of first-time invitees was 50 years old (IQI: 50-50), 15% were immigrants and 33% rarely used the primary healthcare system (Supplementary Table 2). Before the pandemic, 53% of first-time invitees participated in screening within 90 days, 54% within 180 days and 55% within 365 days (data not shown). A slight reduction in participation within 90 days was observed during pre-lockdown (PR=0.95; 95% CI: 0.93-0.98), an increase in participation was found during 1^st^ lockdown (PR=1.06; 95% CI: 1.03-1.09) whereas the participation was similar to the previous years for the remaining part of the study period (Supplementary Table 3).

### Compliance with colonoscopy during the COVID-19 pandemic

There were 94,373 positive FIT tests (in 92,848 individuals) during the study period. Among those 53.7% were men, the median age was 65 years old (IQI=57-70) and 93.3% were of Danish descent (Table 3). Before the pandemic, 89.9% had a colonoscopy performed within 60 days since a positive FIT test (Supplementary Tables 4). The results were unchanged when extending the length of follow-up time to 365 days (data not shown). A minor reduction in compliance with colonoscopy within 60 days was seen during 1^st^ lockdown (Figure 2) reflected in a prevalence ratio (PR) of 0.96 (95% CI: 0.93-0.98) (Table 4). The reduction corresponded to a compliance rate of 85.2% (Supplementary Table 4). The compliance remained stable throughout the rest of the study period (Table 4).

**Table 3.** Baseline characteristics of people with a positive FIT test from colorectal cancer screening in Denmark from 2018 to 2021.

|  | Pre-pandemic<br>(01Jan2018-31Jan2020) | Pre-lockdown<br>(01Feb2020-10Mar2020) | 1st lockdown<br>(11Mar2020-15Apr2020) | 1st re-opening<br>(16Apr2020-15Dec2020) | 2nd lockdown<br>(16Dec2020-27Feb2021) | 2nd re-opening<br>(28Feb2021-30Sep2021) | Total |
| --- | --- | --- | --- | --- | --- | --- | --- |
|  | N (%) | N (%) | N (%) | N (%) | N (%) | N (%) | N (%) |
| **Total** | 54886 (58.2) | 2942 (3.1) | 1319 (1.4) | 16628 (17.6) | 5383 (5.7) | 13215 (14.0) | 94373 (100.0) |
| **Sex** |  |  |  |  |  |  |  |
| Men | 29880 (54.4) | 1532 (52.1) | 704 (53.4) | 8796 (52.9) | 2835 (52.7) | 6918 (52.3) | 50665 (53.7) |
| Women | 25006 (45.6) | 1410 (47.9) | 615 (46.6) | 7832 (47.1) | 2548 (47.3) | 6297 (47.7) | 43708 (46.3) |
| **Age at invitation** |  |  |  |  |  |  |  |
| 50-54 years | 9481 (17.3) | 291 (9.9) | 185 (14.0) | 2615 (15.7) | 987 (18.3) | 2876 (21.8) | 16435 (17.4) |
| 55-59 years | 8606 (15.7) | 838 (28.5) | 157 (11.9) | 3536 (21.3) | 813 (15.1) | 1479 (11.2) | 15429 (16.3) |
| 60-64 years | 10343 (18.8) | 524 (17.8) | 78 (5.9) | 2944 (17.7) | 1000 (18.6) | 2433 (18.4) | 17322 (18.4) |
| 65-69 years | 12126 (22.1) | 559 (19.0) | 111 (8.4) | 3526 (21.2) | 1165 (21.6) | 2888 (21.9) | 20375 (21.6) |
| 70-74 years | 14330 (26.1) | 730 (24.8) | 788 (59.7) | 4007 (24.1) | 1418 (26.3) | 3539 (26.8) | 24812 (26.3) |
| Median (IQI) | 64 (57; 70) | 63 (56; 69) | 74 (59; 75) | 63 (56; 69) | 64 (57; 70) | 64 (57; 70) | 64 (57; 70) |
| **Ethnicity** |  |  |  |  |  |  |  |
| Danish descent | 51287 (93.6) | 2735 (93.0) | 1221 (92.6) | 15484 (93.1) | 4716 (92.4) | 11344 (92.5) | 86787 (93.3) |
| Western Immigrant | 1306 (2.4) | 66 (2.2) | 37 (2.8) | 388 (2.3) | 130 (2.5) | 302 (2.5) | 2229 (2.4) |
| Non-western immigrant | 2205 (4.0) | 140 (4.8) | 61 (4.6) | 756 (4.5) | 257 (5.0) | 620 (5.1) | 4039 (4.3) |
| **Cohabitation status** |  |  |  |  |  |  |  |
| Single | 16451 (30.0) | 939 (31.9) | 463 (35.1) | 5046 (30.3) | 1578 (30.9) | 3575 (29.1) | 28052 (30.1) |
| Co-habiting/co-living | 4273 (7.8) | 231 (7.9) | 88 (6.7) | 1377 (8.3) | 431 (8.4) | 1037 (8.5) | 7437 (8.0) |
| Married/registered partner | 34074 (62.2) | 1771 (60.2) | 768 (58.2) | 10205 (61.4) | 3094 (60.6) | 7654 (62.4) | 57566 (61.9) |
| **Educational level (ISCED)** |  |  |  |  |  |  |  |
| ISCED15 level 1-2 | 27676 (51.3) | 1577 (54.6) | 605 (46.6) | 8988 (54.9) | 2823 (53.6) | 7064 (54.4) | 48733 (52.5) |
| ISCED15 level 3-4 | 20618 (38.2) | 1048 (36.3) | 521 (40.1) | 5741 (35.1) | 1907 (36.2) | 4682 (36.1) | 34517 (37.2) |
| ISCED15 level 5-8 | 5666 (10.5) | 264 (9.1) | 172 (13.3) | 1632 (10.0) | 540 (10.2) | 1231 (9.5) | 9505 (10.2) |
| **Disposable income** |  |  |  |  |  |  |  |
| Lowest quintile | 11851 (21.6) | 556 (18.9) | 321 (24.3) | 3124 (18.8) | 962 (17.9) | 2481 (18.8) | 19295 (20.5) |
| Second quintile | 12530 (22.8) | 690 (23.5) | 351 (26.6) | 3555 (21.4) | 1211 (22.5) | 2954 (22.4) | 21291 (22.6) |
| Third quintile | 11077 (20.2) | 608 (20.7) | 234 (17.7) | 3439 (20.7) | 1042 (19.4) | 2479 (18.8) | 18879 (20.0) |
| Fourth quintile | 10142 (18.5) | 569 (19.4) | 210 (15.9) | 3321 (20.0) | 1046 (19.5) | 2615 (19.9) | 17903 (19.0) |
| Highest quintile | 9244 (16.9) | 517 (17.6) | 203 (15.4) | 3165 (19.1) | 1114 (20.7) | 2641 (20.1) | 16884 (17.9) |
| **Healthcare usage** |  |  |  |  |  |  |  |
| Rare | 8431 (15.4) | 409 (13.9) | 161 (12.2) | 2576 (15.5) | 770 (14.3) | 1903 (14.4) | 14250 (15.1) |
| Low | 8450 (15.4) | 440 (15.0) | 171 (13.0) | 2628 (15.8) | 833 (15.5) | 1957 (14.8) | 14479 (15.3) |
| Average | 11964 (21.8) | 608 (20.7) | 260 (19.7) | 3704 (22.3) | 1205 (22.4) | 2823 (21.4) | 20564 (21.8) |
| High | 11300 (20.6) | 673 (22.9) | 269 (20.4) | 3502 (21.1) | 1126 (20.9) | 2921 (22.1) | 19791 (21.0) |
| Frequent | 14741 (26.9) | 812 (27.6) | 458 (34.7) | 4218 (25.4) | 1449 (26.9) | 3611 (27.3) | 25289 (26.8) |
| **Time from positive FIT to colonoscopy, median (IDI)** | 13 (8; 29) | 13 (7; 31) | 13 (8; 31) | 13 (8; 29) | 13 (8; 28) | 13 (8; 29) | 13 (8; 29) |

IQI = interquartile interval; IDI = interdecile interval; ISCED = International Standard Classification of Education

**Table 4.** Prevalence ratios (PR) and 95% confidence intervals (CI) of compliance with colonoscopy within 60 days since a positive FIT test from colorectal cancer screening in Denmark from 2018-2021.

|  |  | **Pre-pandemic**<br>(01Jan2018-31Jan2020) |  | **Pre-lockdown**<br>(01Feb2020-10Mar2020) |  | **1st lockdown**<br>(11Mar2020-15Apr2020) |  | **1st re-opening**<br>(16Apr2020-15Dec2020) |  | **2nd lockdown**<br>(16Dec2020-27Feb2021) |  | **2nd re-opening**<br>(28Feb2021-30Sep2021) |  |
| --- | --- | --- | --- | --- | --- | --- | --- | --- | --- | --- | --- | --- | --- |
|  |  | N=54982 |  | N=2944 |  | N=1319 |  | N=16628 |  | N=5390 |  | N=13222 |  |
|  | N | PR | [95%CI] | PR | [95%CI] | PR | [95%CI] | PR | [95%CI] | PR | [95%CI] | PR | [95%CI] |
| **Overall** | 94373 | 1.00 | - | 0.98 | [0.96; 1.00] | 0.96 | [0.93; 0.98] | 0.99 | [0.98; 1.00] | 1.01 | [1.00; 1.03] | 0.99 | [0.98; 1.01] |
| **Sex** |  |  |  |  |  |  |  |  |  |  |  |  |  |
| Men | 50665 | 1.00 | - | 0.97 | [0.95; 0.99] | 0.94 | [0.91; 0.97] | 0.99 | [0.98; 0.99] | 0.99 | [0.97; 1.00] | 0.97 | [0.96; 0.98] |
| Women | 43708 | 1.00 | - | 0.98 | [0.96; 1.00] | 0.96 | [0.93; 0.99] | 0.99 | [0.98; 1.00] | 1.00 | [0.99; 1.02] | 0.99 | [0.98; 1.00] |
| **Age at invitation** |  |  |  |  |  |  |  |  |  |  |  |  |  |
| 50-54 years | 16435 | 1.00 | - | 1.04 | [1.00; 1.08] | 0.98 | [0.93; 1.04] | 1.00 | [0.98; 1.03] | 1.01 | [0.97; 1.05] | 0.98 | [0.94; 1.01] |
| 55-59 years | 15429 | 1.00 | - | 0.97 | [0.94; 1.01] | 0.94 | [0.87; 1.01] | 1.00 | [0.98; 1.02] | 1.02 | [0.98; 1.05] | 1.02 | [0.98; 1.06] |
| 60-64 years | 17322 | 1.00 | - | 1.00 | [0.97; 1.04] | 1.04 | [0.98; 1.11] | 0.99 | [0.97; 1.01] | 1.00 | [0.97; 1.04] | 0.98 | [0.94; 1.01] |
| 65-69 years | 20375 | 1.00 | - | 0.99 | [0.96; 1.03] | 1.04 | [0.98; 1.10] | 0.99 | [0.97; 1.01] | 1.03 | [1.00; 1.06] | 1.00 | [0.97; 1.03] |
| 70-74 years | 24812 | 2.00 | - | 0.94 | [0.91; 0.98] | 0.94 | [0.90; 0.97] | 0.98 | [0.96; 1.00] | 1.01 | [0.98; 1.04] | 0.99 | [0.96; 1.02] |
| **Ethnicity** |  |  |  |  |  |  |  |  |  |  |  |  |  |
| Danish descent | 86787 | 1.00 | - | 0.99 | [0.97; 1.00] | 0.96 | [0.94; 0.99] | 1.00 | [0.99; 1.01] | 1.02 | [1.00; 1.03] | 1.00 | [0.98; 1.01] |
| Western immigrant | 2229 | 1.00 | - | 0.87 | [0.75; 1.02] | 0.84 | [0.68; 1.04] | 0.97 | [0.90; 1.05] | 0.99 | [0.87; 1.13] | 0.99 | [0.89; 1.11] |
| Non-western immigrant | 4039 | 1.00 | - | 0.91 | [0.82; 1.01] | 0.90 | [0.78; 1.05] | 0.98 | [0.92; 1.04] | 1.00 | [0.92; 1.10] | 0.95 | [0.87; 1.04] |
| **Cohabitation status** |  |  |  |  |  |  |  |  |  |  |  |  |  |
| Single | 28052 | 1.00 | - | 0.97 | [0.93; 1.00] | 0.95 | [0.90; 1.00] | 1.00 | [0.97; 1.02] | 1.04 | [1.01; 1.08] | 1.04 | [1.01; 1.08] |
| Cohabiting | 7437 | 1.00 | - | 1.02 | [0.96; 1.08] | 1.01 | [0.93; 1.09] | 1.01 | [0.98; 1.05] | 1.03 | [0.98; 1.08] | 1.03 | [0.98; 1.08] |
| Married | 57566 | 1.00 | - | 0.99 | [0.97; 1.00] | 0.96 | [0.93; 0.99] | 0.99 | [0.98; 1.00] | 1.00 | [0.98; 1.02] | 1.00 | [0.98; 1.02] |
| **Educational level (ISCED)** |  |  |  |  |  |  |  |  |  |  |  |  |  |
| ISCED15 level 1-2 | 48733 | 1.00 | - | 1.00 | [0.98; 1.02] | 0.94 | [0.91; 0.98] | 0.99 | [0.97; 1.00] | 1.01 | [0.99; 1.03] | 0.98 | [0.96; 1.00] |
| ISCED15 level 3-4 | 34517 | 1.00 | - | 0.97 | [0.95; 1.00] | 0.97 | [0.94; 1.01] | 1.00 | [0.99; 1.02] | 1.01 | [0.99; 1.04] | 1.01 | [0.98; 1.03] |
| ISCED15 level 5-8 | 9505 | 1.00 | - | 0.94 | [0.89; 1.00] | 0.97 | [0.91; 1.03] | 0.99 | [0.96; 1.02] | 1.02 | [0.98; 1.07] | 0.99 | [0.94; 1.03] |
| **Disposable income** |  |  |  |  |  |  |  |  |  |  |  |  |  |
| Lowest quintile | 19295 | 1.00 | - | 0.96 | [0.92; 1.00] | 0.91 | [0.86; 0.96] | 0.99 | [0.96; 1.01] | 1.02 | [0.98; 1.06] | 0.99 | [0.95; 1.02] |
| Second quintile | 21291 | 1.00 | - | 0.98 | [0.94; 1.02] | 0.92 | [0.87; 0.98] | 1.01 | [0.99; 1.04] | 1.04 | [1.01; 1.08] | 1.02 | [0.99; 1.06] |
| Third quintile | 18879 | 1.00 | - | 0.99 | [0.95; 1.02] | 0.98 | [0.93; 1.03] | 0.98 | [0.96; 1.00] | 0.99 | [0.96; 1.03] | 0.99 | [0.96; 1.02] |
| Fourth quintile | 17903 | 1.00 | - | 0.97 | [0.94; 1.00] | 0.98 | [0.93; 1.02] | 0.99 | [0.97; 1.01] | 1.00 | [0.97; 1.03] | 0.98 | [0.95; 1.01] |
| Highest quintile | 16884 | 1.00 | - | 1.01 | [0.98; 1.05] | 1.02 | [0.97; 1.06] | 0.99 | [0.97; 1.01] | 1.00 | [0.97; 1.03] | 0.98 | [0.95; 1.00] |
| **Healthcare usage** |  |  |  |  |  |  |  |  |  |  |  |  |  |
| Rare | 14250 | 1.00 | - | 0.98 | [0.94; 1.02] | 0.98 | [0.92; 1.04] | 1.01 | [0.99; 1.04] | 1.03 | [0.99; 1.07] | 0.99 | [0.96; 1.03] |
| Low | 14479 | 1.00 | - | 0.97 | [0.93; 1.01] | 0.97 | [0.92; 1.03] | 0.99 | [0.97; 1.02] | 0.99 | [0.95; 1.02] | 0.98 | [0.95; 1.01] |
| Average | 20564 | 1.00 | - | 1.01 | [0.98; 1.04] | 0.94 | [0.89; 0.99] | 1.00 | [0.98; 1.02] | 1.02 | [0.99; 1.05] | 0.99 | [0.96; 1.02] |
| High | 19791 | 1.00 | - | 0.99 | [0.95; 1.02] | 0.97 | [0.92; 1.01] | 0.98 | [0.96; 1.01] | 1.01 | [0.97; 1.04] | 1.01 | [0.97; 1.04] |
| Frequent | 25289 | 1.00 | - | 0.96 | [0.93; 1.00] | 0.95 | [0.90; 0.99] | 0.98 | [0.96; 1.01] | 1.01 | [0.98; 1.04] | 0.98 | [0.95; 1.01] |

\* Adjusted for year, month and age at invitation; PR=prevalence ratio; CI=confidence interval; ISCED=International Standard Classification of Education

**Figure 2.**
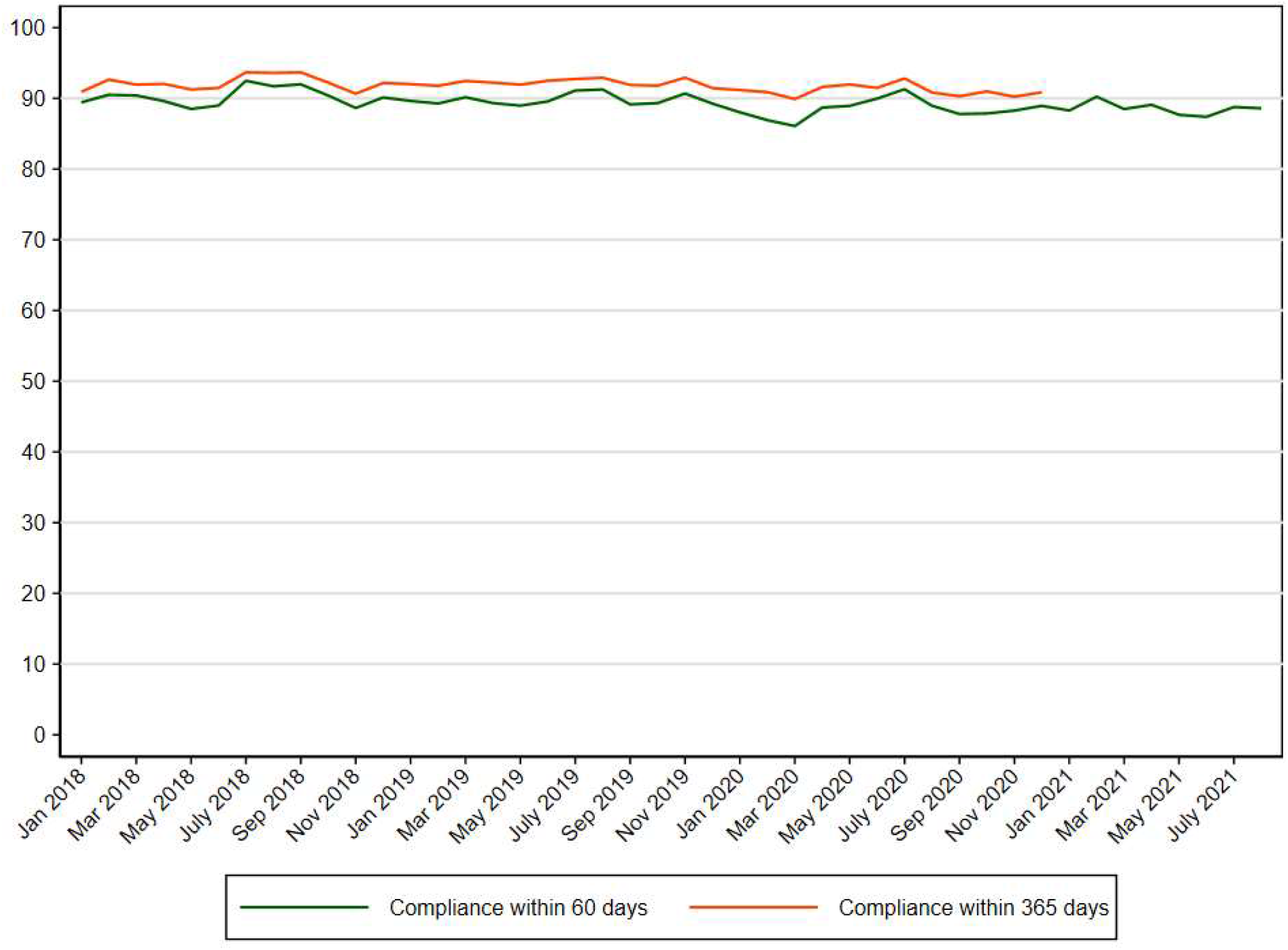
Compliance with colonoscopy (%) within 60 and 365 days since a positive FIT test from colorectal cancer screening in Denmark from 2018-2021.

Immigrants, individuals living alone and individuals with a low income had a lower compliance with colonoscopy before the pandemic (Supplementary Figure 3). During pre-lockdown and 1^st^ lockdown, the compliance with colonoscopy within 60 days was lower among both 55-59 year olds and 70-74 year olds, among immigrants and among individuals with a low income compared with the previous years (Table 4). The results were unchanged when extending the length of follow-up time to 365 days (data not shown).

### Time to participation

Before the pandemic, the median time from invitation to participation was 28 days (IDI=16-72) increasing to 32 days (IDI=16-64) during 1^st^ lockdown and returning to 28 days during 1^st^ reopening (Table 1).

### Sensitivity analysis

When conducting a sensitivity analysis accounting for the reduction in the number of invitations in week 11-14 2020, the estimates were almost identical (data not shown).

## DISCUSSION

### Main findings

In this nation-wide population-based study comprising more than 3,1 million invitations (in 1,928,725 individuals), we found that 60.7% participated in colorectal cancer screening within 90 days before the pandemic. A minor reduction in participation was observed at the start of the pandemic corresponding to a participation rate of 54.9% during pre-lockdown and 53.0% during 1st lockdown. From the 1^st^ re-opening of the society and onwards a relative 5-10% increased participation in screening was seen corresponding to a participation rate of up to 64.9%. The largest relative increase in participation was observed among 55-59 year olds, individuals living alone or cohabiting and immigrants. Among 94,373 positive FIT tests (in 92,848 individuals), we saw that the compliance with colonoscopy within 60 days was 89.9% before the pandemic. A slight reduction was observed during 1^st^ lockdown, where after it resumed to normal levels.

### Comparison with previous studies

In most of the world, the colorectal cancer screening programmes were paused at the start of the pandemic, whilst Denmark was one of the only countries to have the programme running throughout the pandemic. The situation in Denmark is therefore unique and resembles “a natural experiment” illustrating what happens if a screening programme based on faecal samples obtained at home is kept open during a pandemic.

In Spain, the colorectal cancer screening programme was paused for the first seven months of the pandemic leading to a reduced participation in stool-based screening and subsequent colonoscopy (4). The screening programme in Spain (4) requires the invitees to collect a FIT test kit at a pharmacy, which could cause an additional barrier to participation in particular during a pandemic. The programme in Denmark is on the contrary based on a home-based test mailed directly to the invitees. Also in England, the colorectal cancer screening programme based on faecal samples was paused at the start of the pandemic, which led to large reductions in the number of people referred, diagnosed, and treated for colorectal cancer at the start of the pandemic (1). In the Netherlands, the colorectal cancer screening programme was temporarily halted at the start of the pandemic resulting in a lower participation in FIT screening and fewer colorectal cancers diagnosed (2). In Canada, the FIT-based screening programme was suspended at the start of the pandemic resulting in a large reduction in the colorectal cancer faecal test volume (5). A closure of the screening programme may therefore have large detrimental effects on the diagnosing and treatment of colorectal cancer. We found a minor reduction in participation at the start of the pandemic corresponding to a participation rate of 54.9% during pre-lockdown and 53.0% during 1st lockdown indicating that a few people do not participate at the very early phase of a pandemic. These results did not differ when extending the length of follow-up time.

A Danish study by Skovlund et al. found a 24% reduction in the number of colon cancers diagnosed from April to June 2020 in Denmark (7) and a proportion of those may in theory have occurred as a result of the reduction in screening participation. However, as only 20% of all colorectal cancers are diagnosed by screening in Denmark (22), the minor reduction in screening participation that we found during the first two months of the pandemic in Denmark is unlikely to explain the reduction in colon cancer as reported by Skovlund et al. Thus the reduction in the number of colon cancers diagnosed at the start of the pandemic may be caused either by a change in health-seeking behaviour with fever referred to colonoscopy after symptoms, a reduction of coincidental detections of colorectal cancers as a result of elective surveillance colonoscopy being cancelled or postponed or it may be caused by delayed registration of cancers at the time of the study by Skovlund et al.

We found an overall 5-10% increased participation in colorectal cancer screening from 1^st^ reopening of the society and during the subsequent lockdowns. The initial colorectal cancer screening test is a home-based FIT test and people may have had more time to go through their mail and to reach a decision to participate in colorectal cancer screening during the pandemic because many people continued to work from home during different phases of the pandemic; however, on the contrary, we saw a slight reduction in participation at the start of the pandemic. Another possible explanation may be that the pandemic led to a greater degree of health awareness leading to an increased participation in screening. Nonetheless, in the other two screening programmes (breast and cervical cancer screening), we did not find an increased participation during 1^st^ re-opening and the subsequent lockdowns (23, 24), most likely because those programmes require that the invitees visit their general practitioner or a mammography screening clinic.

The study from Spain (4) found a reduction in participation in subsequent colonoscopy as a result of the screening programme being closed during the first seven months of the pandemic. The study from England also showed a marked reduction in the number of colonoscopies (1) as a result of the screening programme being paused at the start of the pandemic. The study from the Netherlands showed a reduced participation in follow-up colonoscopy in the months before and during the suspension of the FIT screening programme (3). We found a 4% reduction in compliance with colonoscopy within 60 days during 1^st^ lockdown despite the programme being open. Fortunately, the compliance with colonoscopy resumed to the same level as before the pandemic from 1^st^ reopening and onwards. Congruently, a qualitative study from the United Kingdom (25) found that interview participants were concerned about visiting healthcare settings at the start of the pandemic, which could explain the slightly lower compliance with colonoscopy.

### Socio-economic differences

In line with previous studies (8, 9), we found that the participation in colorectal cancer screening was lowest among the youngest age group, among men, among immigrants, among individuals living alone or cohabiting, among individuals with a low educational level, a low income and among individuals who rarely use the healthcare system. We found an overall 5-10% increased participation in screening from 1^st^ re-opening and onwards. The largest relatively increases in participation was observed among individuals aged 55-59 years old, among individuals living alone or cohabiting and among immigrants; however, those groups are also the ones who had the lowest participation before the pandemic and a small percentage increase will result in a large prevalence ratio. Possibly, the effect of working from home on screening participation is most pronounced among 55-59 year olds who are still on the labour market, whilst the older age groups may be gradually leaving the labour market, which could explain the increased participation in this particular age group. A qualitative study from Denmark found that immigrants generally have a mistrust in the Danish healthcare system and e.g., prefer a second opinion in their native country (26). However, during the pandemic immigrants may not have been able to travel to their home country and may therefore have opted to participate in screening in Denmark instead of in their home-country.

Furthermore, in accordance with previous studies (9), we found that immigrants, people living alone and people with a low income had a lower compliance with colonoscopy. The reduction in compliance at the start of the pandemic was most pronounced among 55-59 year olds, 70-74 year olds, among immigrants and among persons with a low income. The oldest age group may be reluctant to come into contact with the healthcare system amidst a pandemic, immigrants may not understand the importance of participating in screening as shown in the qualitative study from Denmark (26) and may not understand the information conveyed at the national televised press conferences during the COVID-19 pandemic if that information is only provided in Danish. Other factors may also act as barriers for persons with a low income to participate in colonoscopy e.g., being able to take time off work to participate in colonoscopy and reluctance to use public transport during a pandemic may also act as a barrier.

### Strengths and limitations

We used high-quality population-based data covering the Danish population invited to participate in colorectal cancer screening, which is a major strength of the study. The completeness of the Danish registries is high (27), which also confers to the Danish Colorectal Cancer Screening Database (20). Limitations of the study should also be acknowledged. We did not have data on underlying disease, which may affect individuals’ participation in colorectal cancer screening and compliance with colonoscopy. We did; however, include age which is strongly associated with the level of comorbidity and we thereby reduce the theoretical impact of comorbidity on the results.

Additionally, we did not have data on COVID-19 vaccination status, which may affect colorectal cancer screening participation and compliance with colonoscopy.

### Implications

The initial colorectal cancer screening test is a home-based test and does thereby not require contact with the healthcare system – or use of e.g., public transport to reach a healthcare facility. In theory, this screening modality should be little affected by the pandemic; nonetheless, a slight reduction in participation was observed at the start of the pandemic perhaps because people counterbalance the importance of screening participation amidst a pandemic as seen in other screening programmes (28). It is therefore important to ensure that the health communication is clear and that people are made aware that the colorectal cancer screening is open and that people can safely participate in both the initial screening test and in the subsequent colonoscopy.

We found an overall 5-10% increased participation in colorectal cancer screening from 1^st^ reopening and onwards. As a result of the work-from-home mandate people spent more time at home which may have given people more time to participate in screening. We did not find an increased participation in the other cancer screening programmes (breast and cervical cancer) in Denmark (23, 24) most likely because those programmes require contact with the healthcare system. A homebased screening test thus appear to work well during a pandemic, which could be used in other programmes e.g., cervical cancer screening.

The overall compliance with colonoscopy was largely unaffected by the pandemic, which gives reassurance that individuals in need of follow-up because of a positive FIT test do attend colonoscopy amidst a pandemic. Nonetheless, we found that the compliance with colonoscopy was reduced among 55-59 year olds, 70-74 year olds, among immigrants and among persons with a low income indicating that some groups are affected adversely by the pandemic. Worryingly, some cancers may have gone undetected or diagnosed at a later stage in these groups of individuals.

## Conclusion

In this nation-wide study, we found that the participation in the Danish FIT-based colorectal cancer screening programme and subsequent compliance to colonoscopy after a positive FIT test was only slightly affected by the COVID-19 pandemic.

## Data Availability

Data availability statement In order to comply with the Danish regulations on data privacy, the datasets generated and analysed during this project are not publicly available as the data are stored and maintained electronically at Statistics Denmark, where it only can be accessed by pre-approved researchers using a secure VPN remote access. Furthermore, no data at a personal level nor data not exclusively necessary for publication are allowed to be extracted from the secure data environment at Statistics Denmark. Access to the data can however be granted by the authors of the present study upon a reasonable scientific proposal within the boundaries of the present project and for scientific purposes only.

## Authorship

1. TBO, HJ, HM, JWJ, BA and MR designed study. TBO and HJ acquired the data. HJ analysed the data. All authors contributed to the interpretation of the data.
2. TBO drafted the article. All authors revised the article critically for important intellectual content.
3. All authors approved the final version of the article to be published.
4. All authors agree to be accountable for all aspects of the work in ensuring that questions related to the accuracy or integrity of any part of the work are appropriately investigated and resolved.

## Materials availability statement

Not applicable.

## Data availability statement

In order to comply with the Danish regulations on data privacy, the datasets generated and analysed during this project are not publicly available as the data are stored and maintained electronically at Statistics Denmark, where it only can be accessed by pre-approved researchers using a secure VPN remote access. Furthermore, no data at a personal level nor data not exclusively necessary for publication are allowed to be extracted from the secure data environment at Statistics Denmark. Access to the data can however be granted by the authors of the present study upon a reasonable scientific proposal within the boundaries of the present project and for scientific purposes only.

## SUPPLEMENTARY TABLES AND FIGURES

**Supplementary Figure 1.**
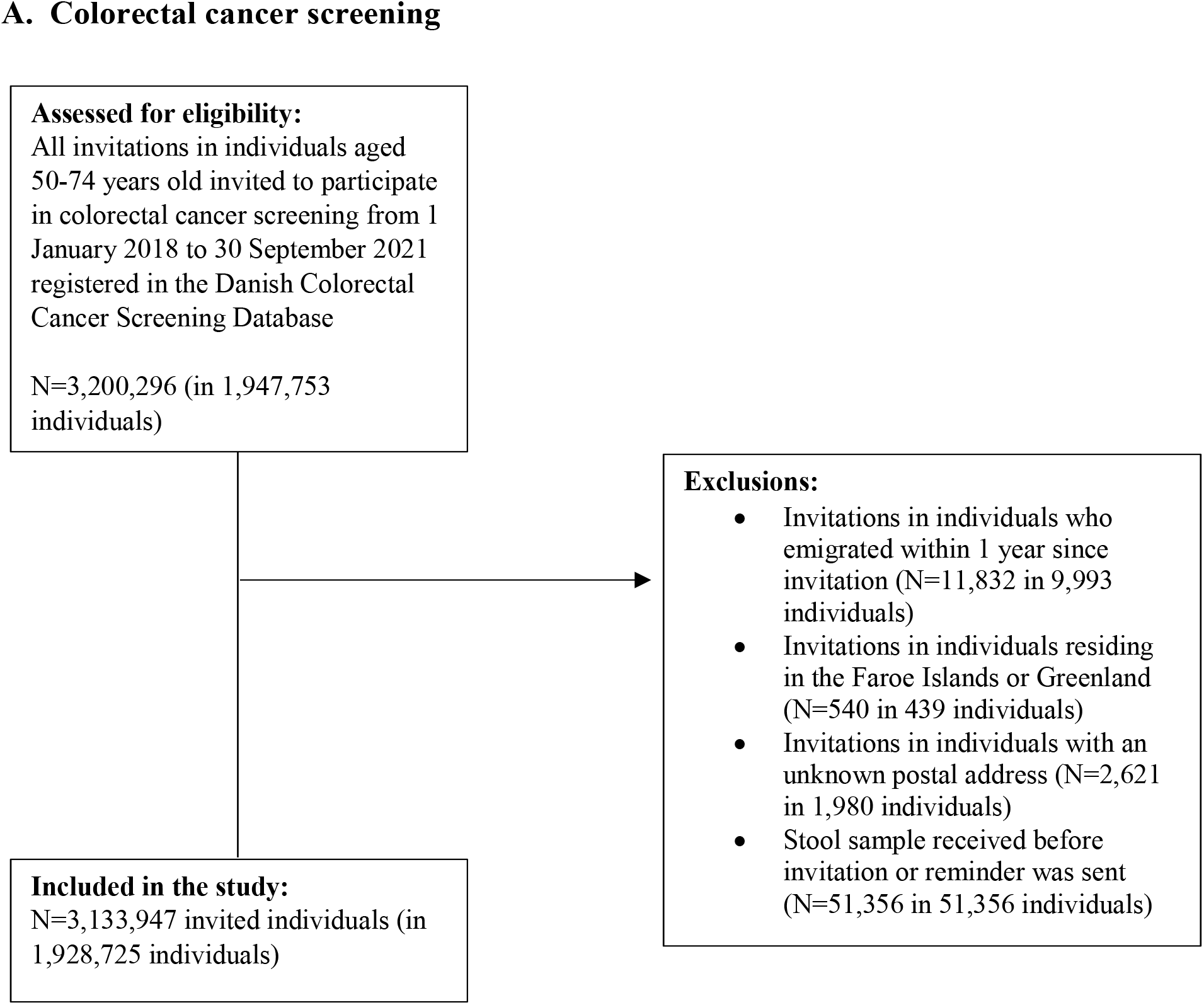
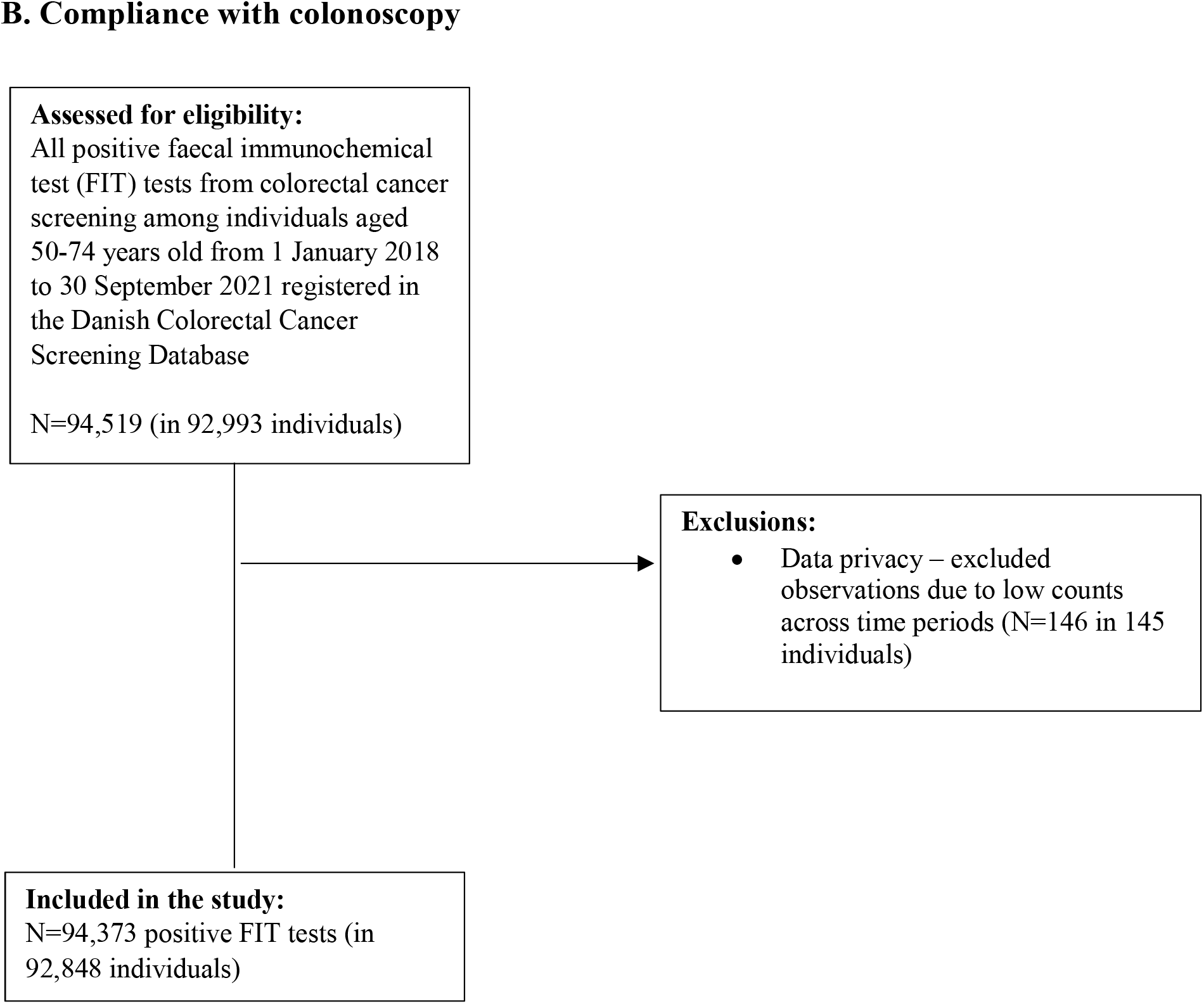
Flow-chart of the study population.

**Supplementary Table 1.** Participation in colorectal cancer screening (%) in Denmark within 90 days since invitation from 2018 to 2021.

|  | **Pre-pandemic**<br>**(01Jan2018-**<br>**31Jan2020)** | **Pre-lockdown**<br>**(01Feb2020-**<br>**10Mar2020)** | **1st lockdown**<br>**(11Mar2020-**<br>**15Apr2020)** | **1st re-opening**<br>**(16Apr2020-**<br>**15Dec2020)** | **2nd lockdown**<br>**(16Dec2020-**<br>**27Feb2021)** | **2nd re-opening**<br>**(28Feb2021-**<br>**30Sep2021)** |
| --- | --- | --- | --- | --- | --- | --- |
|  | % | % | % | % | % | % |
| **Total** | 60.7 | 54.9 | 53.0 | 62.4 | 63.0 | 64.9 |
| **Age at invitation** |  |  |  |  |  |  |
| 50-54 years | 54.3 | 45.2 | 56.0 | 51.8 | 53.2 | 56.7 |
| 55-59 years | 57.2 | 50.2 | 60.1 | 62.3 | 64.7 | 64.4 |
| 60-64 years | 62.2 | 56.6 | 61.6 | 63.9 | 63.9 | 66.0 |
| 65-69 years | 67.2 | 62.7 | 64.7 | 68.5 | 69.0 | 70.8 |
| 70-74 years | 66.7 | 65.0 | 47.2 | 69.6 | 69.9 | 72.3 |
| **Ethnicity** |  |  |  |  |  |  |
| Danish descent | 61.6 | 55.7 | 53.5 | 63.2 | 64.3 | 66.8 |
| Descendant of immigrant | 54.5 | 50.3 | 44.6 | 56.9 | 58.1 | 60.8 |
| Western Immigrant | 53.5 | 47.6 | 47.6 | 54.2 | 57.9 | 59.3 |
| Non-western immigrant | 50.8 | 45.5 | 47.1 | 54.1 | 56.2 | 57.9 |
| **Cohabitation status** |  |  |  |  |  |  |
| Single | 49.5 | 43.6 | 43.0 | 51.1 | 51.8 | 54.8 |
| Co-habiting/co-living | 55.3 | 49.9 | 52.9 | 57.2 | 58.8 | 61.1 |
| Married/registered partner | 67.5 | 62.0 | 59.3 | 69.4 | 70.6 | 72.4 |
| **Educational level (ISCED)** |  |  |  |  |  |  |
| ISCED15 level 1-2 | 58.2 | 52.5 | 52.8 | 60.3 | 60.4 | 62.8 |
| ISCED15 level 3-4 | 64.1 | 58.5 | 53.0 | 65.7 | 66.7 | 68.3 |
| ISCED15 level 5-8 | 65.1 | 58.9 | 55.2 | 66.9 | 68.3 | 69.1 |
| **Disposable income** |  |  |  |  |  |  |
| Lowest quintile | 54.7 | 49.5 | 46.1 | 55.7 | 56.6 | 59.0 |
| Second quintile | 56.7 | 51.5 | 46.2 | 57.9 | 58.1 | 60.6 |
| Third quintile | 61.9 | 55.8 | 55.0 | 63.1 | 63.0 | 65.5 |
| Fourth quintile | 65.0 | 58.9 | 60.7 | 66.6 | 66.8 | 68.2 |
| Highest quintile | 65.0 | 57.9 | 63.1 | 67.3 | 68.2 | 69.2 |
| **Healthcare usage** |  |  |  |  |  |  |
| Rare | 49.5 | 42.9 | 44.9 | 51.1 | 51.0 | 52.4 |
| Low | 60.0 | 54.5 | 54.2 | 61.9 | 62.3 | 63.8 |
| Average | 64.0 | 58.2 | 56.0 | 66.0 | 66.4 | 68.3 |
| High | 65.7 | 60.6 | 55.9 | 67.5 | 68.2 | 70.4 |
| Frequent | 65.3 | 60.4 | 53.1 | 67.2 | 68.0 | 70.3 |

ISCED=International Standard Classification of Education

**Supplementary Figure 2.**
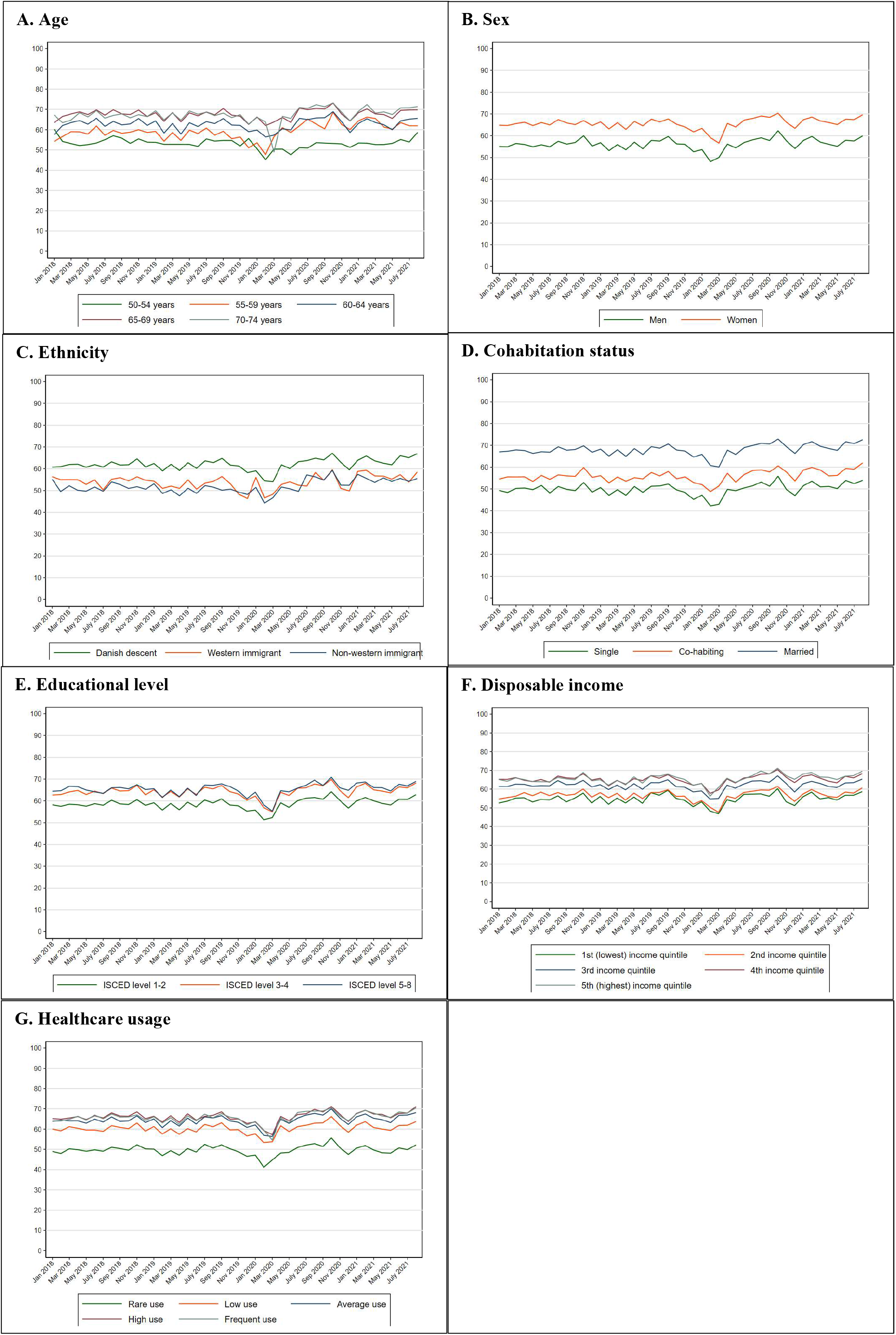
Participation in colorectal cancer screening (%) in Denmark within 90 days since invitation from 2018 to 2021 stratified by the explanatory variables.

**Supplementary Table 2.** Baseline characteristics of first-time invitees invited to participate in colorectal cancer screening in Denmark 2018-2021.

|  | Pre-pandemic<br>(01Jan2018-31Jan2020) | Pre-lockdown<br>(01Feb2020-10Mar2020) | 1st lockdown<br>(11Mar2020-15Apr2020) | 1st re-opening<br>(16Apr2020-15Dec2020) | 2nd lockdown<br>(16Dec2020-27Feb2021) | 2nd re-opening<br>(28Feb2021-30Sep2021) | Total |
| --- | --- | --- | --- | --- | --- | --- | --- |
|  | N (%) | N (%) | N (%) | N (%) | N (%) | N (%) | N (%) |
| **Total** | 148704 (53.8) | 8278 (3.0) | 6779 (2.5) | 54328 (19.6) | 14358 (5.2) | 44048 (15.9) | 276495 (100.0) |
| **Sex** |  |  |  |  |  |  |  |
| Men | 75340 (50.7) | 4188 (50.6) | 3327 (49.1) | 27269 (50.2) | 7303 (50.9) | 22161 (50.3) | 139588 (50.5) |
| Women | 73364 (49.3) | 4090 (49.4) | 3452 (50.9) | 27059 (49.8) | 7055 (49.1) | 21887 (49.7) | 136907 (49.5) |
| **Age at invitation** |  |  |  |  |  |  |  |
| 50-54 years | 144126 (96.9) | 8034 (97.1) | 6673 (98.4) | 52708 (97.0) | 13903 (96.8) | 42728 (97.0) | 268172 (97.0) |
| 55-59 years | 1817 (1.2) | 108 (1.3) | 13 (0.2) | 663 (1.2) | 180 (1.3) | 570 (1.3) | 3351 (1.2) |
| 60-64 years | 1221 (0.8) | 62 (0.7) | 5 (0.1) | 436 (0.8) | 118 (0.8) | 317 (0.7) | 2159 (0.8) |
| 65-69 years | 882 (0.6) | 47 (0.6) | 12 (0.2) | 306 (0.6) | 88 (0.6) | 260 (0.6) | 1595 (0.6) |
| 70-74 years | 658 (0.4) | 27 (0.3) | 76 (1.1) | 215 (0.4) | 69 (0.5) | 173 (0.4) | 1218 (0.4) |
| Median (IQR) | 50 (50; 50) | 50 (50; 50) | 50 (50; 50) | 50 (50; 50) | 50 (50; 50) | 50 (50; 50) | 50 (50; 50) |
| **Ethnicity** |  |  |  |  |  |  |  |
| Danish descent | 127132 (85.7) | 6941 (84.1) | 5567 (82.3) | 45931 (84.7) | 2540 (84.2) | N/A | 188111 (85.3) |
| Western Immigrant | 6920 (4.7) | 481 (5.8) | 279 (4.1) | 2913 (5.4) | 157 (5.2) | N/A | 10750 (4.9) |
| Non-western immigrant | 14023 (9.5) | 812 (9.8) | 903 (13.3) | 5254 (9.7) | 311 (10.3) | N/A | 21303 (9.7) |
| **Cohabitation status** |  |  |  |  |  |  |  |
| Single | 47755 (32.2) | 2660 (32.2) | 2181 (32.2) | 17518 (32.3) | 949 (31.5) | N/A | 71063 (32.2) |
| Co-habiting/co-living | 19413 (13.1) | 1074 (13.0) | 857 (12.7) | 7263 (13.4) | 408 (13.5) | N/A | 29015 (13.2) |
| Married/registered partner | 81146 (54.7) | 4518 (54.8) | 3729 (55.1) | 29469 (54.3) | 1658 (55.0) | N/A | 120520 (54.6) |
| **Educational level (ISCED)** |  |  |  |  |  |  |  |
| ISCED15 level 1-2 | 93443 (64.0) | 5229 (64.1) | 4316 (64.9) | 34039 (64.2) | 9030 (65.3) | 27169 (64.5) | 173226 (64.2) |
| ISCED15 level 3-4 | 37534 (25.7) | 2046 (25.1) | 1676 (25.2) | 13166 (24.8) | 3408 (24.6) | 10521 (25.0) | 68351 (25.3) |
| ISCED15 level 5-8 | 15086 (10.3) | 885 (10.8) | 659 (9.9) | 5827 (11.0) | 1393 (10.1) | 4440 (10.5) | 28290 (10.5) |
| **Disposable income** |  |  |  |  |  |  |  |
| Lowest quintile | 21311 (14.4) | 1015 (12.6) | 901 (13.4) | 6618 (12.5) | 1737 (12.4) | 4817 (11.3) | 36399 (13.4) |
| Second quintile | 18355 (12.4) | 962 (11.9) | 918 (13.6) | 6109 (11.6) | 1567 (11.1) | 4554 (10.7) | 32465 (11.9) |
| Third quintile | 29964 (20.2) | 1601 (19.8) | 1260 (18.7) | 10453 (19.8) | 2484 (17.7) | 7164 (16.8) | 52926 (19.4) |
| Fourth quintile | 37041 (25.0) | 2049 (25.4) | 1720 (25.5) | 13595 (25.8) | 3516 (25.0) | 10949 (25.7) | 68870 (25.3) |
| Highest quintile | 41551 (28.0) | 2439 (30.2) | 1946 (28.9) | 15997 (30.3) | 4757 (33.8) | 15162 (35.6) | 81852 (30.0) |
| **Healthcare usage** |  |  |  |  |  |  |  |
| Rare | 45423 (30.5) | 2947 (35.6) | 2287 (33.7) | 19872 (36.6) | 5191 (36.2) | 15825 (35.9) | 91545 (33.1) |
| Low | 32255 (21.7) | 1512 (18.3) | 1281 (18.9) | 10297 (19.0) | 2755 (19.2) | 8314 (18.9) | 56414 (20.4) |
| Average | 31566 (21.2) | 1657 (20.0) | 1366 (20.2) | 10473 (19.3) | 2789 (19.4) | 8525 (19.4) | 56376 (20.4) |
| High | 21804 (14.7) | 1154 (13.9) | 987 (14.6) | 7458 (13.7) | 2002 (13.9) | 6137 (13.9) | 39542 (14.3) |
| Frequent | 17656 (11.9) | 1008 (12.2) | 858 (12.7) | 6228 (11.5) | 1621 (11.3) | 5247 (11.9) | 32618 (11.8) |
| **Time - invitation to participation, median (IDI)** | 38 (17; 92) | 44 (16; 107) | 41 (17; 86) | 38 (17; 87) | 35 (17; 81) | 31 (17; 74) | 37 (17; 88) |

IQI = interquartile interval; IDI = interdecile interval; ISCED = International Standard Classification of Education

**Supplementary Table 3.** Prevalence ratios (PR) and 95% confidence intervals (CI) of participation in colorectal cancer screening within 90 days since invitation in Denmark among first-time invitees 2018-2021*.

|  |  | Pre-pandemic<br>(01Jan2018-31Jan2020)<br>N=148704 |  | Pre-lockdown<br>(01Feb2020-10Mar2020)<br>N=8278 |  | 1st lockdown<br>(11Mar2020-15Apr2020)<br>N=6779 |  | 1st re-opening<br>(16Apr2020-15Dec2020)<br>N=54328 |  | 2nd lockdown<br>(16Dec2020-27Feb2021)<br>N=14358 |  | 2nd re-opening<br>(28Feb2021-30Sep2021)<br>N=44048 |  |
| --- | --- | --- | --- | --- | --- | --- | --- | --- | --- | --- | --- | --- | --- |
|  | N | PR | [95%CI] | PR | [95%CI] | PR | [95%CI] | PR | [95%CI] | PR | [95%CI] | PR | [95%CI] |
| **Overall** | 276495 | 1.00 | - | 0.95 | [0.93; 0.98] | 1.06 | [1.03; 1.09] | 0.98 | [0.97; 1.00] | 1.03 | [1.01; 1.06] | 1.01 | [0.98; 1.03] |
| **Sex** |  |  |  |  |  |  |  |  |  |  |  |  |  |
| Men | 139588 | 1.00 | - | 0.92 | [0.89; 0.96] | 1.08 | [1.05; 1.12] | 1.00 | [0.99; 1.02] | 1.04 | [1.02; 1.07] | 1.03 | [1.01; 1.04] |
| Women | 136907 | 1.00 | - | 0.95 | [0.92; 0.98] | 1.03 | [1.00; 1.06] | 1.00 | [0.98; 1.01] | 1.01 | [0.99; 1.03] | 1.01 | [1.00; 1.02] |
| **Age at invitation** |  |  |  |  |  |  |  |  |  |  |  |  |  |
| 50-54 years | 268172 | 1.00 | - | 0.96 | [0.93; 0.98] | 1.06 | [1.03; 1.09] | 0.99 | [0.97; 1.01] | 1.04 | [1.01; 1.06] | 1.02 | [0.99; 1.04] |
| 55-59 years | 3351 | 1.00 | - | 0.76 | [0.52; 1.11] | 0.45 | [0.12; 1.69] | 0.94 | [0.75; 1.16] | 1.16 | [0.83; 1.61] | 1.05 | [0.77; 1.43] |
| 60-64 years | 2159 | 1.00 | - | 0.83 | [0.57; 1.20] | 0.97 | [0.32; 2.98] | 0.85 | [0.68; 1.07] | 0.79 | [0.57; 1.11] | 0.79 | [0.57; 1.08] |
| 65-69 years | 1595 | 1.00 | - | 0.96 | [0.65; 1.41] | 0.60 | [0.26; 1.39] | 0.96 | [0.75; 1.22] | 0.94 | [0.66; 1.36] | 0.95 | [0.67; 1.34] |
| 70-74 years | 1218 | 1.00 | - | 1.05 | [0.67; 1.64] | 1.03 | [0.70; 1.52] | 1.04 | [0.79; 1.37] | 0.88 | [0.60; 1.30] | 0.84 | [0.57; 1.24] |
| **Ethnicity** |  |  |  |  |  |  |  |  |  |  |  |  |  |
| Danish descent | 188111 | 1.00 | - | 0.96 | [0.93; 0.99] | 1.05 | [1.02; 1.08] | 0.98 | [0.96; 1.00] | 1.01 | [0.97; 1.05] | N/A | N/A |
| Western Immigrant | 10750 | 1.00 | - | 1.01 | [0.86; 1.18] | 1.11 | [0.95; 1.31] | 1.03 | [0.93; 1.14] | 1.14 | [0.93; 1.39] | N/A | N/A |
| Non-western immigrant | 21303 | 1.00 | - | 0.94 | [0.85; 1.05] | 1.10 | [1.00; 1.21] | 1.01 | [0.94; 1.07] | 1.09 | [0.96; 1.24] | N/A | N/A |
| **Cohabitation status** |  |  |  |  |  |  |  |  |  |  |  |  |  |
| Single | 71063 | 1.00 | - | 0.94 | [0.88; 1.00] | 1.05 | [0.99; 1.11] | 0.95 | [0.92; 0.99] | 0.98 | [0.90; 1.06] | N/A | N/A |
| Cohabiting | 29015 | 1.00 | - | 1.02 | [0.93; 1.11] | 1.09 | [1.00; 1.18] | 1.00 | [0.95; 1.06] | 1.03 | [0.92; 1.15] | N/A | N/A |
| Married | 120520 | 1.00 | - | 0.96 | [0.92; 0.99] | 1.05 | [1.02; 1.09] | 1.00 | [0.97; 1.02] | 1.03 | [0.99; 1.08] | N/A | N/A |
| **Educational level (ISCED)** |  |  |  |  |  |  |  |  |  |  |  |  |  |
| ISCED15 level 1-2 | 173226 | 1.00 | - | 0.96 | [0.93; 0.99] | 1.06 | [1.02; 1.10] | 0.99 | [0.97; 1.01] | 1.04 | [1.01; 1.07] | 1.02 | [0.98; 1.05] |
| ISCED15 level 3-4 | 68351 | 1.00 | - | 0.92 | [0.87; 0.97] | 1.04 | [0.99; 1.10] | 0.97 | [0.93; 1.00] | 1.03 | [0.98; 1.08] | 0.99 | [0.94; 1.04] |
| ISCED15 level 5-8 | 28290 | 1.00 | - | 0.98 | [0.90; 1.06] | 1.12 | [1.03; 1.21] | 1.02 | [0.97; 1.08] | 1.03 | [0.95; 1.11] | 1.06 | [0.98; 1.15] |
| **Disposable income** |  |  |  |  |  |  |  |  |  |  |  |  |  |
| Lowest quintile | 36399 | 1.00 | - | 1.00 | [0.90; 1.11] | 1.16 | [1.05; 1.29] | 1.01 | [0.95; 1.07] | 1.09 | [0.99; 1.19] | 1.02 | [0.93; 1.12] |
| Second quintile | 32465 | 1.00 | - | 1.01 | [0.92; 1.11] | 1.07 | [0.98; 1.18] | 1.02 | [0.96; 1.08] | 1.05 | [0.96; 1.14] | 1.09 | [1.00; 1.19] |
| Third quintile | 52926 | 1.00 | - | 0.98 | [0.92; 1.04] | 1.09 | [1.03; 1.16] | 1.00 | [0.96; 1.04] | 1.03 | [0.97; 1.09] | 1.02 | [0.96; 1.08] |
| Fourth quintile | 68870 | 1.00 | - | 0.94 | [0.89; 0.98] | 1.05 | [1.00; 1.10] | 0.98 | [0.95; 1.01] | 1.00 | [0.95; 1.04] | 0.98 | [0.94; 1.03] |
| Highest quintile | 81852 | 1.00 | - | 0.94 | [0.90; 0.98] | 1.04 | [0.99; 1.09] | 0.99 | [0.97; 1.02] | 1.04 | [1.00; 1.08] | 1.01 | [0.97; 1.05] |
| **Healthcare usage** |  |  |  |  |  |  |  |  |  |  |  |  |  |
| Rare | 91545 | 1.00 | - | 0.92 | [0.87; 0.97] | 1.04 | [0.98; 1.09] | 0.96 | [0.92; 0.99] | 1.01 | [0.96; 1.07] | 0.96 | [0.91; 1.01] |
| Low | 56414 | 1.00 | - | 0.99 | [0.93; 1.05] | 1.13 | [1.06; 1.19] | 1.01 | [0.98; 1.05] | 1.05 | [0.99; 1.10] | 1.06 | [1.01; 1.12] |
| Average | 56376 | 1.00 | - | 0.95 | [0.90; 1.01] | 1.03 | [0.97; 1.08] | 0.99 | [0.95; 1.02] | 1.01 | [0.96; 1.07] | 0.98 | [0.93; 1.03] |
| High | 39542 | 1.00 | - | 0.99 | [0.92; 1.06] | 1.06 | [0.99; 1.13] | 1.01 | [0.97; 1.05] | 1.04 | [0.98; 1.10] | 1.06 | [0.99; 1.12] |
| Frequent | 32618 | 1.00 | - | 0.97 | [0.90; 1.04] | 1.09 | [1.02; 1.17] | 0.99 | [0.95; 1.04] | 1.08 | [1.01; 1.15] | 1.00 | [0.94; 1.07] |

\* Adjusted for year, month and age at invitation; PR=prevalence ratio; CI=confidence interval; ISCED=International Standard Classification of Education

**Supplementary Table 4.** Compliance with colonoscopy (%) in Denmark within 60 days since a positive FIT test from 2018 to 2021.

|  | Pre-pandemic<br>(01Jan2018-31Jan2020) | Pre-lockdown<br>(01Feb2020-10Mar2020) | 1st lockdown<br>(11Mar2020-15Apr2020) | 1st re-opening<br>(16Apr2020-15Dec2020) | 2nd lockdown<br>(16Dec2020-27Feb2021) | 2nd re-opening<br>(28Feb2021-30Sep2021) |
| --- | --- | --- | --- | --- | --- | --- |
|  | % | % | % | % | % | % |
| **Total** | 89.9 | 87.6 | 85.2 | 88.8 | 89.4 | 88.2 |
| **Age at invitation** |  |  |  |  |  |  |
| 50-54 years | 89.9 | 92.8 | 89.2 | 90.2 | 89.4 | 87.9 |
| 55-59 years | 90.3 | 87.1 | 82.8 | 88.9 | 89.7 | 89.3 |
| 60-64 years | 90.2 | 89.7 | 93.6 | 89.4 | 89.8 | 88.0 |
| 65-69 years | 90.2 | 88.4 | 91.9 | 88.4 | 90.0 | 87.9 |
| 70-74 years | 89.3 | 84.0 | 83.0 | 87.7 | 88.4 | 88.2 |
| **Ethnicity** |  |  |  |  |  |  |
| Danish descent | 90.3 | 88.3 | 86.0 | 89.2 | 89.9 | 88.6 |
| Western Immigrant | 86.3 | 75.8 | 73.0 | 82.7 | 83.1 | 84.4 |
| Non-western immigrant | 83.9 | 79.3 | 77.0 | 84.0 | 86.8 | 81.8 |
| **Cohabitation status** |  |  |  |  |  |  |
| Single | 85.7 | 81.5 | 79.5 | 83.8 | 85.9 | 83.9 |
| Co-habiting/co-living | 90.4 | 89.6 | 88.6 | 90.8 | 90.3 | 87.2 |
| Married/registered partner | 92.0 | 90.6 | 88.3 | 91.0 | 91.3 | 90.3 |
| **Educational level (ISCED)** |  |  |  |  |  |  |
| ISCED15 level 1-2 | 89.4 | 88.6 | 83.6 | 88.3 | 89.3 | 87.5 |
| ISCED15 level 3-4 | 90.5 | 87.7 | 86.8 | 89.7 | 89.8 | 89.4 |
| ISCED15 level 5-8 | 91.0 | 85.2 | 88.4 | 89.5 | 90.6 | 89.2 |
| **Disposable income** |  |  |  |  |  |  |
| Lowest quintile | 87.7 | 83.6 | 81.3 | 85.6 | 87.4 | 85.5 |
| Second quintile | 87.7 | 83.3 | 79.2 | 86.2 | 87.2 | 85.3 |
| Third quintile | 90.4 | 88.8 | 87.6 | 88.6 | 88.0 | 89.0 |
| Fourth quintile | 92.3 | 90.2 | 91.0 | 91.2 | 92.5 | 90.7 |
| Highest quintile | 92.6 | 93.2 | 93.1 | 92.5 | 91.8 | 90.6 |
| **Healthcare usage** |  |  |  |  |  |  |
| Rare | 90.8 | 88.0 | 87.0 | 90.3 | 90.1 | 87.4 |
| Low | 91.2 | 88.2 | 88.9 | 90.9 | 90.0 | 90.2 |
| Average | 91.2 | 91.0 | 84.6 | 90.0 | 90.7 | 89.0 |
| High | 90.2 | 88.6 | 87.0 | 87.8 | 88.5 | 89.2 |
| Frequent | 87.4 | 83.7 | 82.5 | 86.3 | 88.2 | 86.0 |

ISCED = International Standard Classification of Education

**Supplementary Figure 3.**
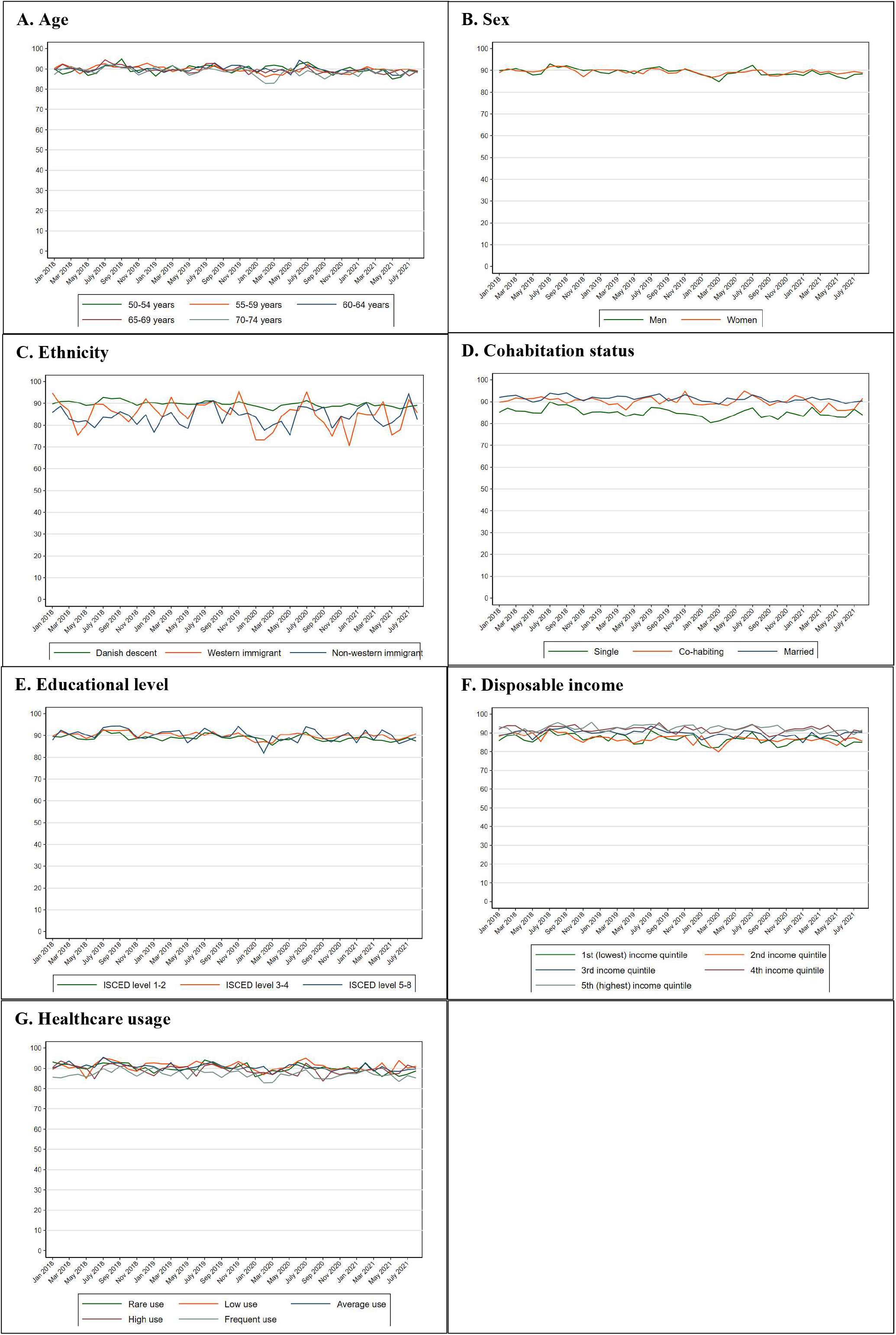
Compliance with colonoscopy (%) in Denmark within 60 days since a positive FIT test from 2018 to 2021 stratified by the explanatory variables.

